# A Clinically-Informed Framework for Evaluating Vision-Language Models in Radiology Report Generation: Taxonomy of Errors and Risk-Aware Metric

**DOI:** 10.1101/2025.07.13.25331222

**Authors:** Hao Guan, Peter C. Hou, Pengyu Hong, Liqin Wang, Wenyu Zhang, Xinsong Du, Zhengyang Zhou, Li Zhou

## Abstract

Recent advances in vision-language models (VLMs) have enabled automatic radiology report generation, yet current evaluation methods remain limited to general-purpose NLP metrics or coarse classification-based clinical scores. In this study, we propose a clinically informed evaluation framework for VLM-generated radiology reports that goes beyond traditional performance measures. We define a taxonomy of 12 radiology-specific error types, each annotated with clinical risk levels (low, medium, high) in collaboration with physicians. Using this framework, we conduct a comprehensive error analysis of three representative VLMs, i.e., DeepSeek VL2, CXR-LLaVA, and CheXagent, on 685 gold-standard, expert-annotated MIMIC-CXR cases. We further introduce a risk-aware evaluation metric, the Clinical Risk-weighted Error Score for Text-generation (CREST), to quantify safety impact. Our findings reveal critical model vulnerabilities, common error patterns, and condition-specific risk profiles, offering actionable insights for model development and deployment. This work establishes a safety-centric foundation for evaluating and improving medical report generation models. The source code of our evaluation framework, including CREST computation and error taxonomy analysis, is available at https://github.com/guanharry/VLM-CREST.

## Introduction

The integration of Vision-Language Models (VLMs) into radiology report generation offers promising potential to streamline diagnostic workflows and reduce radiologists’ workloads.^1,2,3,4^ By combining chest X-ray interpretation with natural language generation, VLMs can produce reports that support clinical decision-making and save time.^5^ However, VLM-generated reports are still prone to serious mistakes, such as hallucinations (fabricated findings), omissions (missed abnormalities), and incorrect expressions of uncertainty. These errors can undermine clinical trust, threaten patient safety, and ultimately impede real-world deployment.

Despite recent progress, most prior work on VLMs in radiology has focused on general language metrics (e.g., BLEU, ROUGE) or proxy clinical classification scores,^6,7^ which fail to capture the clinical impact of specific errors. Few studies have examined the types of mistakes VLMs make, their clinical severity, or compared how different models fail. The lack of clinically grounded evaluation limits our understanding of VLM safety and reliability in high-stakes environments such as clinical decision-making.

To address these gaps, we propose a clinically informed evaluation framework for assessing errors in VLM-generated radiology reports. We define a taxonomy of 12 clinically relevant error types and assign each a clinical risk level (low, medium, high) in collaboration with physicians. Using this framework, we analyze errors made by three open-source representative VLMs on 685 expert-annotated chest X-ray cases. We introduce a novel **Clinical Risk-weighted Error Score for Text-generation (CREST)** to quantify safety-sensitive model performance and explore the distribution of errors by model, condition, and risk category. Our results reveal critical failure modes, high-risk patterns, and design opportunities for safer, more trustworthy VLMs in clinical radiology applications.

## Methods

### Data Collection

This study uses the publicly available MIMIC-CXR dataset,^8,9^ a large-scale de-identified chest radiograph dataset collected from the Beth Israel Deaconess Medical Center in Boston, MA. The dataset includes frontal and lateral chest X-ray images along with corresponding free-text radiology reports, and is widely used for developing and evaluating AI systems in medical imaging. Please note that the MIMIC-CXR dataset includes an official split of training, validation, and test sets as defined by the dataset creators. All pretrained models used in this study were trained only on the training and validation sets. This ensures that our evaluation on the test set avoids any data leakage and provides a fair assessment of model generalization.

For this study, we selected a subset of 685 cases from MIMIC-CXR (test set) to serve as our **gold-standard evaluation set**. Each case was independently reviewed and annotated by a board-certified radiologist with eight years of clinical experience. This expert-annotated set provides reference for error analysis and evaluation of VLM-generated reports.

Each case was labeled for the presence or absence of 13 common radiological findings, comprising 12 pathological conditions and one non-pathological category:

- **Pathological conditions**: Atelectasis, Cardiomegaly, Consolidation, Edema, Enlarged Cardiomediastinum, Fracture, Lung Lesion, Lung Opacity, Pleural Effusion, Pleural Other, Pneumonia, and Pneumothorax.
- **Support Devices**: While not a disease, this category reflects the presence of medical hardware (e.g. endotracheal tubes) that is clinically important to capture in automated reports.

Each condition was labeled using a four-level scheme:

- **1**: Positive (finding present)
- **0**: Negative (finding explicitly ruled out)
- **-1**: Uncertain (finding mentioned with ambiguity)
- **-2**: Not Mentioned (no reference in the report)

Note: In the original dataset, conditions that are not mentioned in the report are represented as null values. For ease of analysis, we explicitly assign a value of “ -2” to indicate “ Not Mentioned.” This labeling scheme facilitates detailed comparisons between VLM-generated reports and expert annotations, enabling fine-grained clinical error analysis.

### Model Selection

To evaluate the performance and clinical reliability of current VLMs in radiology report generation, we selected three representative models that span a spectrum of open-source, general-purpose, fine-tuned, and domain-specific architectures: *DeepSeek VL2, CXR-LLaVA*, and *CheXagent*. These models were chosen based on their popularity, diversity in training paradigms, and relevance to the medical imaging domain.

1. **DeepSeek VL2:** A state-of-the-art general-purpose VLM developed for a wide range of vision-language tasks.^10,11^ It is trained on large-scale image-text pairs from the general domain and has demonstrated strong performance in natural image understanding and captioning. However, it has not been specifically adapted for medical imaging, making it a suitable model to assess out-of-domain generalization.
2. **CXR-LLaVA:** A chest X-ray-specific adaptation of the open-source LLaVA model.^12^ It is fine-tuned on medical images (chest X-ray) and radiology reports to improve performance in the medical domain while leveraging the core strengths of the original LLaVA framework.^13^ This model represents a middle ground between general-purpose and domain-specialized systems.
3. **CheXagent:** A domain-specific VLM designed specifically for radiology report generation.^14^ It has been trained on large volumes of chest X-rays and their corresponding reports, integrating expert-guided data processing and clinical priors. CheXagent represents the current state-of-the-art in specialized report generation and interpretation for chest radiography.

To standardize input across all models, we used the following prompt for report generation: *“ You are a radiologist analyzing a chest X-ray image. Please provide the ‘impression’ section for the radiology report, summarizing the key findings in a concise and professional manner*.*”* This prompt encourages models to focus on clinically meaningful content and produce concise diagnostic summaries that follow real-world radiology reporting practices.

All selected models are open-source, which was a key criterion in our selection due to their alignment with medical data privacy and safety requirements. Each model was deployed and executed in a controlled local environment equipped with four NVIDIA L40 GPUs (each with 48 GB memory). All models were downloaded and configured following their official instructions from GitHub or Hugging Face. The implementation pipeline included loading pretrained weights, applying uniform image processing, and capturing the generated reports in a structured format for downstream analysis.

### Evaluation Framework

#### Error Taxonomy

To evaluate model performance in a clinically meaningful way, as shown in Table 1, we define a taxonomy of 12 specific error (discordance) types based on label mismatches between the model-generated report and the gold-standard annotations. These error types capture not only whether the model was incorrect, but *how* it was incorrect and *what clinical consequences* the error might entail.

**Table 1:** Error Taxonomy with Definitions, Risk Levels, and Examples.

| Error Type | GT → AI | Definition | Risk Level | Example |
| --- | --- | --- | --- | --- |
| False Negative (Wrong Negation) | 1 → 0 | Model states a finding is negative when it is present. | High | GT: "Left-sided pneumothorax."<br>AI: "No pneumothorax detected." |
| False Negative (Uncertainty Downgrade) | 1 → -1 | Definite finding downgraded to uncertain. | High | GT: "Moderate pleural effusion."<br>AI: "Possible pleural effusion." |
| Omission (Missed Finding) | 1 → -2 | Present finding is omitted. | High | GT: "Left rib fracture."<br>AI: (No mention) |
| False Positive (Wrong Affirmation) | 0 → 1 | Absent condition incorrectly reported as present. | Medium | GT: "No pneumonia."<br>AI: "Pneumonia detected." |
| False Positive (Uncertainty Introduction) | 0 → -1 | Uncertainty introduced to a negative finding. | Medium | GT: "No pneumothorax."<br>AI: "Possible pneumothorax." |
| Omission (Removed Negative Evidence) | 0 → -2 | Explicit negation is dropped. | Low | GT: "No pneumonia."<br>AI: (No mention) |
| Hallucination (Nonexistent Condition) | -2 → 1 | Condition invented without basis. | High | GT: (No mention)<br>AI: "Pneumonia is present." |
| False Certainty (Wrong Negation) | -2 → 0 | Condition falsely declared negative without GT basis. | Low | GT: (No mention)<br>AI: "No pneumothorax." |
| False Certainty (Uncertainty Introduction) | -2 → -1 | Condition falsely flagged as uncertain. | Medium | GT: (No mention)<br>AI: "Possible pleural effusion." |
| Uncertainty Overconfidence (False Positive) | -1 → 1 | Uncertain finding reported with unjustified certainty. | Medium | GT: "Possible opacity."<br>AI: "Opacity present." |
| Uncertainty Underconfidence (False Negative) | -1 → 0 | Uncertain finding negated. | High | GT: "Possible edema."<br>AI: "No edema." |
| Uncertainty Omission (Dropped Finding) | -1 → -2 | Uncertainty statement omitted entirely. | High | GT: "Possible lung opacity."<br>AI: (No mention) |

Each error is described using a **ground-truth-to-AI transition format (GT***→* **AI)** across four possible label states: **1 = Positive, 0 = Negative, -1 = Uncertain**, and **-2 = Not Mentioned**.

The 12 error types fall into four categories, including: 1) False affirmations or negations (e.g., stating presence or absence incorrectly); 2) Mismanagement of uncertainty (e.g., unjustified upgrades or downgrades); 3) Complete omission of findings or negative evidence; 4) Hallucinations, i.e., fabricating findings that were never present. Each error type is also associated with a **clinical risk level** (High, Medium, Low), reflecting its potential harm in practice. These assignments were defined in consultation with physicians.

#### Clinical Risk Mapping

While previous studies on AI-generated radiology reports have focused on linguistic fidelity or alignment with structured labels, few addressed the *clinical severity* of model errors. In this work, we introduce a clinically informed risk stratification scheme that assigns a risk level to each of the 12 defined error types based on their potential to impact patient outcomes. Table 1 includes the assigned clinical risk level for each error type. These levels are further integrated into our proposed risk-aware evaluation metrics. This risk mapping was developed in close collaboration with physicians from Brigham and Women’s Hospital (BWH) to ensure clinical rigor and real-world relevance. Each error type was reviewed and assigned a risk level, i.e., High, Medium, or Low, based on its likelihood to: 1) Delay diagnosis or treatment; 2) Cause misdiagnosis or unnecessary intervention/follow-up; 3) Undermine clinical trust or compromise patient safety.

For example, a *false negative* error, where the model incorrectly states “ no pneumothorax” when a pneumothorax is actually present, is classified as **high risk**, given the potential for life-threatening consequences if left untreated. *False positive* errors are considered **medium risk**, reflecting the clinical reality that over-treatment is often preferred to under-treatment. In contrast, omission of an explicitly negative finding (e.g., failing to mention “ no pneumothorax” when this is stated in the ground truth) is categorized as **low risk**, as such errors rarely lead to big clinical harm.

This clinical risk dimension adds a critical layer of evaluation that goes beyond correctness. It allows us to ask not only *whether* a model made a mistake, but *how dangerous* that mistake is. By establishing a risk mapping that is both domain-expert-driven and reproducible, our framework provides a blueprint for clinical-grade evaluation of AI systems, enhancing the model interpretability and offering actionable insights for both developers and clinicians.

#### Annotation Protocol

We conducted our evaluation using a gold-standard dataset of 685 chest X-ray cases from MIMIC-CXR (test set), each manually annotated by a board-certified radiologist with eight years of clinical experience. For each case, the corresponding radiology report included structured labels covering 13 clinical conditions. Each condition was annotated with one of four possible values: **1** (positive), **0** (negative), **-1** (uncertain), and **-2** (not mentioned). These expert-generated annotations serve as a trusted reference standard and form the foundation for our error analysis.

To extract condition-level predictions from model-generated reports, we employed CheXbert,^15^ a widely used and state-of-the-art radiology report labeler trained on large corpora of clinical text within the same domain. CheXbert converts free-text radiology reports into structured labels that align with the schema used in our gold-standard dataset, thereby enabling consistent, condition-level comparisons between model outputs and reference annotations. CheXbert has become a standard evaluation tool in recent radiology NLP and VLM research,^16,17,18,19^ particularly valued for its clinical schema alignment and robust performance. Furthermore, its classification outputs have been widely adopted as clinical efficacy metrics to assess the real-world utility of AI-generated reports.^2^ This widespread adoption reflects its practicality, scalability, and reproducibility, especially critical in settings where manual re-annotation of large numbers of reports is infeasible.

While CheXbert is not infallible, its labeling behavior is systematic and has been benchmarked against human expert annotations. As such, even if some label noise exists, it is consistent across models and does not significantly affect comparative analysis. Moreover, our evaluation framework does not rely solely on label matching: all errors are further categorized using our manually curated taxonomy and clinically informed risk schema, developed in collaboration with physicians. This semantic and risk-based layer mitigates isolated labeling noise and ensures that performance comparisons reflect meaningful clinical differences rather than annotation artifacts. Additionally, this layered framework enhances the flexibility and extensibility of our approach. It can be paired with any labeler, human or automated, making it adaptable to future studies and larger-scale evaluations.

#### Risk-Aware Evaluation Metrics

To complement our clinical error taxonomy and risk stratification framework, we propose a set of *risk-aware evaluation metrics* that quantify model performance not only by correctness but also by the potential clinical consequences of errors. These metrics provide a more clinical-informed and safety-sensitive alternative to conventional metrics such as BLEU, ROUGE, or classification accuracy.

Our primary metric is the **Clinical Risk-weighted Error Score for Text-generation (CREST)**, which aggregates model errors by weighting each error according to its clinical risk level, i.e., *High, Medium*, or *Low* as defined in Table 1. Let *ℰ* = *{e*_1_, *e*_2_, …, *e*_*N*_ *}* denote the set of all errors made by a model. Each error *e*_*i*_ is assigned a clinical risk weight *r*(*e*_*i*_) ∈ {1, 2, 3} corresponding to low, medium, and high risk, respectively. The CREST is defined as:

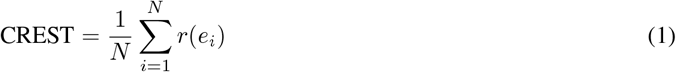

We also define a normalized version to rescale the score between 0 and 1:

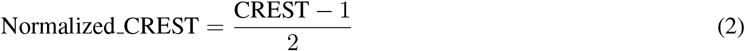

This normalization maps a CREST of 1 (all errors are low-risk) to 0, and a CREST of 3 (all errors are high-risk) to 1. A higher value of CREST indicates more clinically harmful model performance. To further characterize model safety, we introduce a complementary metric:

#### High-Risk Error Rate (HRER)

The proportion of all errors that are high-risk:

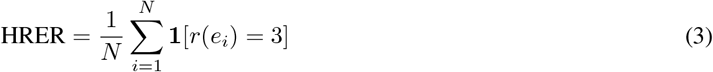

where **1**[*·*] denotes the indicator function (i.e., it equals 1 if the condition is true).

Together, these metrics offer a multidimensional view of model reliability by incorporating clinical risk into performance evaluation. For example, two models may produce the same number of total errors, but one may concentrate its mistakes in low-risk categories, while the other distributes more of its errors in high-risk areas. Such differences are invisible to traditional metrics yet critically important for assessing the real-world safety and clinical applicability of AI systems. By capturing not just the quantity but the potential severity of errors, CREST and its companion metrics provide a principled and safety-aware foundation for AI performance auditing and model comparison.

## Results

### Overall Error Frequency

To establish a baseline for model performance, we begin by analyzing the total number of errors each model made across the 685 gold-standard cases. Each case is annotated with 13 labels corresponding to 13 clinical conditions, resulting in a total of 8,905 labels. Therefore, a model that fails completely would incur 8,905 errors. The distribution of these labels per condition is shown in Figure 1(a).

**Figure 1:**
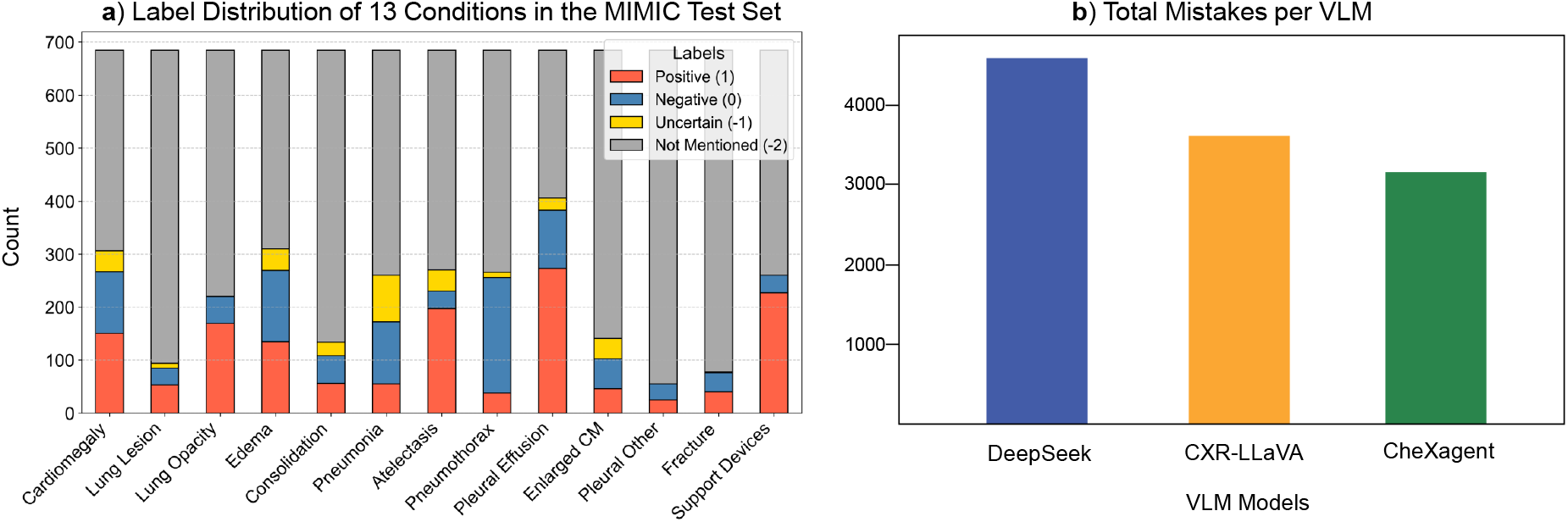
Overall Error Frequency for VLMs.

As shown in Figure 1(b), DeepSeek VL2 produces the highest number of errors, followed by CXR-LLaVA and CheXagent. This aligns with expectations: DeepSeek, a general-purpose model not trained on medical data, struggles more with domain-specific tasks. CXR-LLaVA, fine-tuned on chest X-rays, performs moderately better, while CheXagent, designed specifically for radiology report generation, makes the fewest errors overall.

Although the underlying label distribution is imbalanced, all models were evaluated on the same set of condition labels under consistent assumptions. Therefore, the total error count provides a fair and meaningful comparison of overall model performance. In the following sections, we further examine error types, condition-specific trends, and risk-aware metrics to assess the clinical relevance of these mistakes.

### Error Type Distribution

To better understand the nature of VLM-generated report mistakes, we analyzed the frequency of each specific error type. Rather than relying on raw error counts, which can be biased by imbalanced label distributions, we computed the normalized error rate, defined as the number of errors divided by the total number of corresponding ground-truth labels. For example, consider the error type (1 → -2), which indicates an omission of a definite abnormality. If there are **M** such errors and the total number of ground-truth labels with value 1 is **N**, then the normalized error rate is 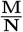. This allows for a fairer comparison across error types.

As shown in Figure 2, the three most frequent normalized error types across all models are: 1) **Uncertainty Omission (-1 → -2)**, 2) **Omission of Negative Evidence (0 → -2)**, and 3) **Missed Positive Findings (1 → -2)**. These errors consistently dominate for all models and highlight a common failure mode: VLMs often omit important clinical information, either by dropping uncertain findings, removing negative evidence, or failing to report definite abnormalities. Such omissions can pose serious clinical risks, especially for high-acuity conditions like pneumothorax, where missed findings may delay critical diagnosis and treatment.

**Figure 2:**
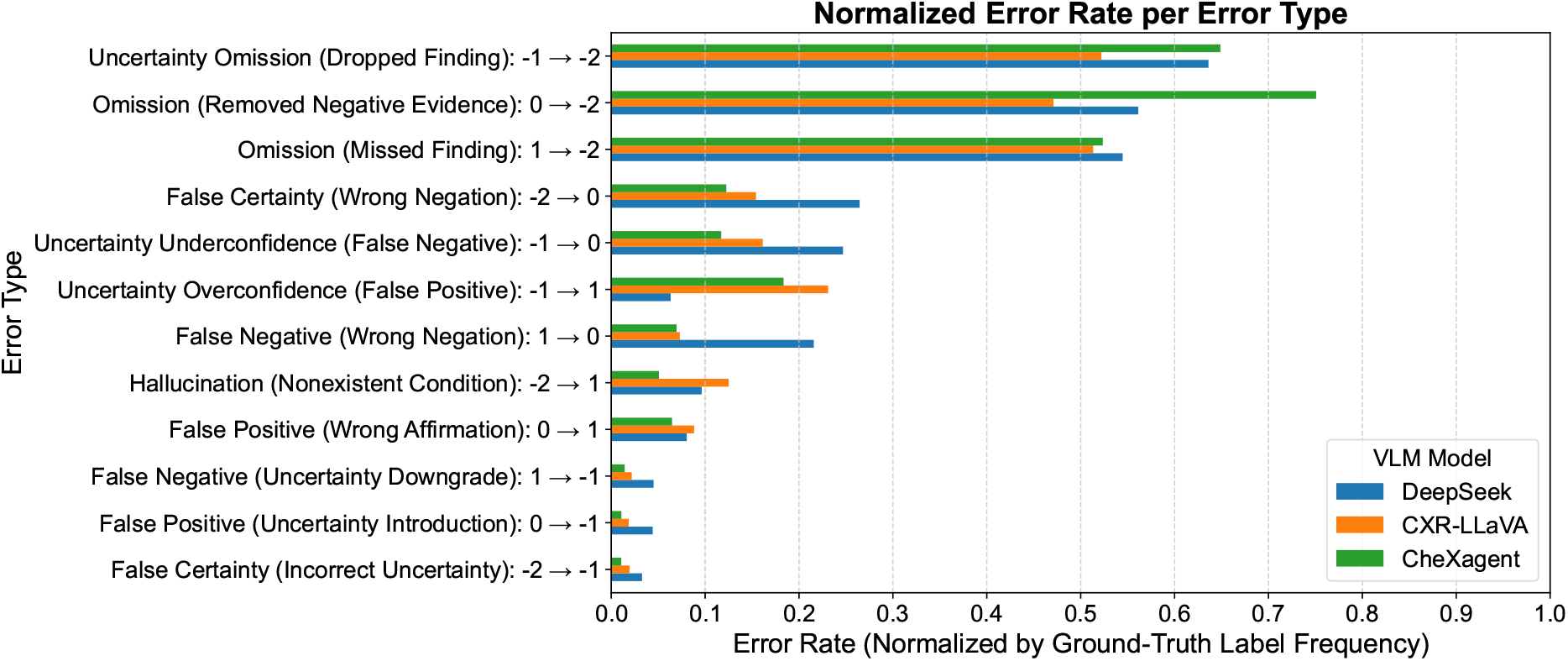
Normalized Error Rate of the Predefined 12 Error Types for Different VLMs.

Interestingly, when accounting for label frequency, errors such as Hallucination (-2 → 1) and False Negation of Uncertainty (-1 → 0) remain relatively moderate, suggesting that overt fabrication or misclassification of uncertainty occurs less frequently relative to the number of corresponding ground-truth instances. In terms of model-specific performance, DeepSeek generally underperforms, as expected for a general-purpose model. However, it does not consistently perform the worst. This highlights that even general-purpose models may exhibit strengths in handling specific types of errors.

### Per-Condition Analysis

To better understand the nature of each model’s failures, we conducted a condition-level breakdown of errors across the 13 clinical conditions. As shown in Figure 3, we report the error rate for each condition, defined as the number of incorrect predictions divided by the total number of ground-truth labels.

**Figure 3:**
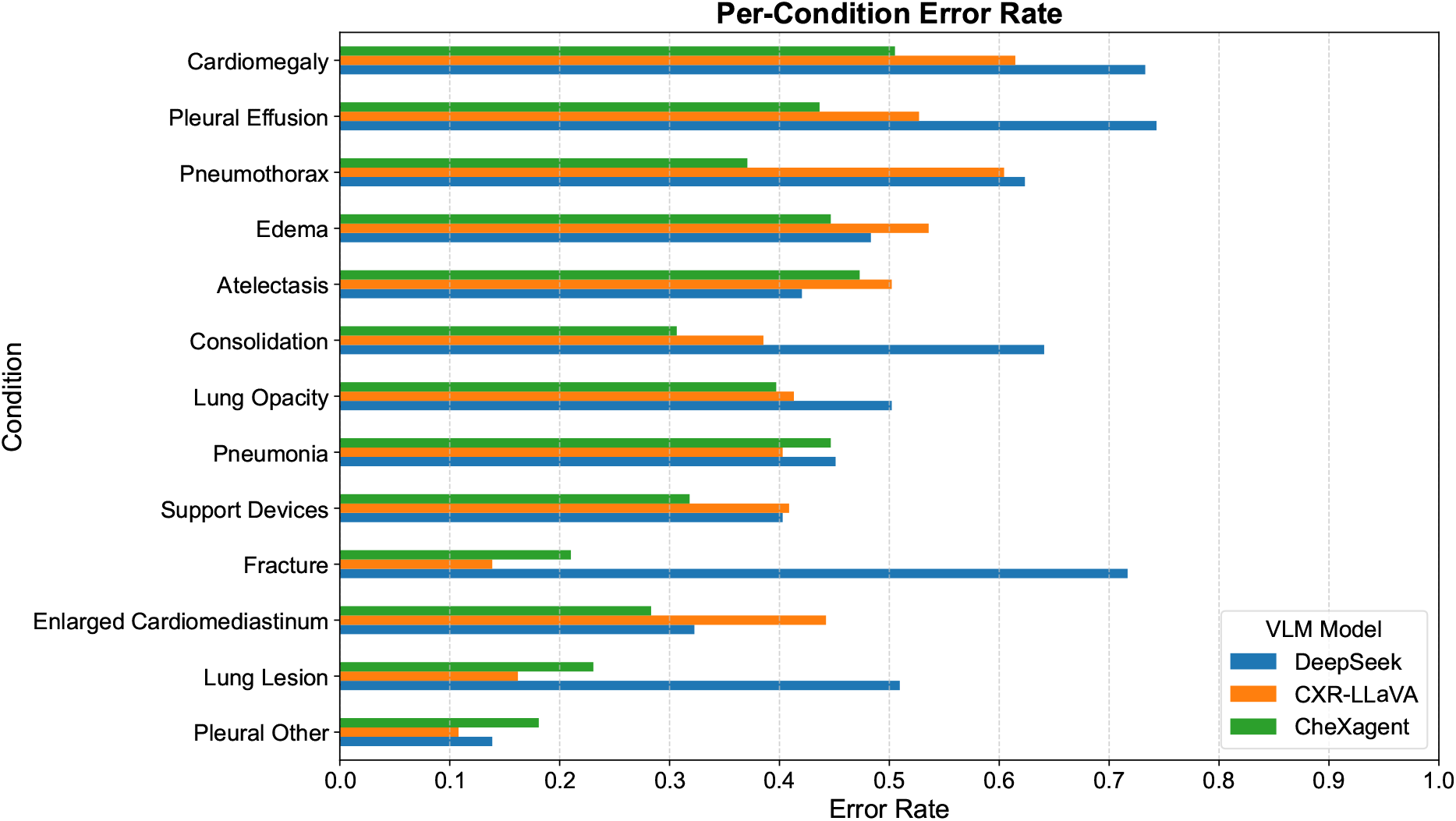
Error Rate for Each Condition across VLMs.

Across nearly all conditions, DeepSeek VL2 shows the highest error rate, which is expected as it is a general-purpose vision-language model not specifically trained on chest X-rays. In contrast, CXR-LLaVA and CheXagent, both adapted or fine-tuned for the radiology domain, consistently produce fewer errors across conditions.

Interestingly, while CXR-LLaVA outperforms DeepSeek in most categories, it shows comparable or higher error rates in some clinically sensitive findings such as Pneumothorax, Atelectasis, and Edema. It suggests that the model, though domain-specialized, may still struggle with subtle or ambiguous presentations of critical findings. CheXagent shows the most consistent performance across conditions, with lower or similar error counts in nearly every category. This reflects its task-specific design and closer alignment with the labeling conventions used in training and evaluation.

This condition-specific breakdown allows us to pinpoint model vulnerabilities. For instance, all models struggle disproportionately with pleural effusion and cardiomegaly, possibly due to their frequent co-occurrence or subtle radiographic manifestations. Such analysis not only reveals which conditions are hardest for current models, but also informs clinical prioritization for future model development and fine-tuning.

### Clinical Risk Distribution

Beyond error counts, understanding the clinical implications of each mistake is crucial for assessing the safety of radiology report generation. Using the expert-defined risk mapping, we categorized each error made by the models into High, Medium, or Low Risk based on its potential clinical harm.

Figure 4 shows the exact numbers and proportion of errors at each risk level for the three evaluated VLMs. The High-Risk Error Rate (HRER) can be clearly observed in this figure. While CXR-LLaVA, a domain-adapted model fine-tuned on chest X-rays, produced fewer overall mistakes than DeepSeek VL2, it exhibited the highest proportion of high-risk errors. This suggests that although CXR-LLaVA better recognizes many visual patterns, its failures, when they occur, tend to involve more clinically critical conditions. In contrast, CheXagent displayed a favorable profile, with the smallest number of high-risk errors and a relatively large portion of low-risk errors, such as missing explicitly negative findings.

**Figure 4:**
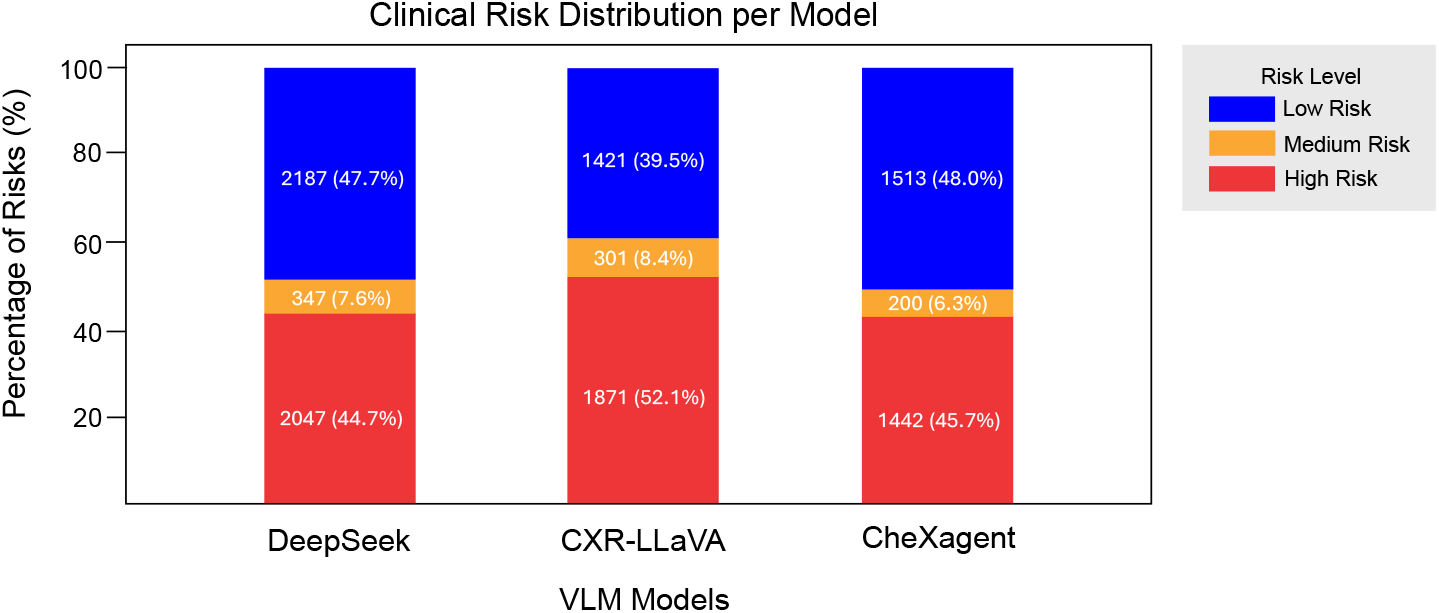
Clinical Risk Distribution.

This finding highlights a trade-off: a model may be more accurate overall, yet more prone to serious errors in high-stakes scenarios. Risk profiling therefore complements traditional metrics by offering a deeper view of model performance that aligns with real-world clinical priorities.

### Clinical Risk-weighted Error Score for Text-generation (CREST)

To assess the clinical severity of errors made by the VLMs, we calculate the normalized CREST scores for each of the 13 clinical conditions. An average normalized CREST score is then computed to serve as an overall indicator of clinical risk.

Figure 5 presents the averaged normalized CREST scores across all 13 clinical conditions for the three VLMs. CXR-LLaVA exhibits the highest score (0.57), indicating it tends to make more clinically severe errors on average, despite its overall lower error count. DeepSeek follows with a score of 0.50, showing moderate risk behavior, while CheXagent achieves the lowest CREST score (0.46), suggesting a more conservative and risk-averse error profile. These results emphasize that models with fewer total mistakes (e.g., CXR-LLaVA) can still exhibit higher clinical risk per error.

**Figure 5:**
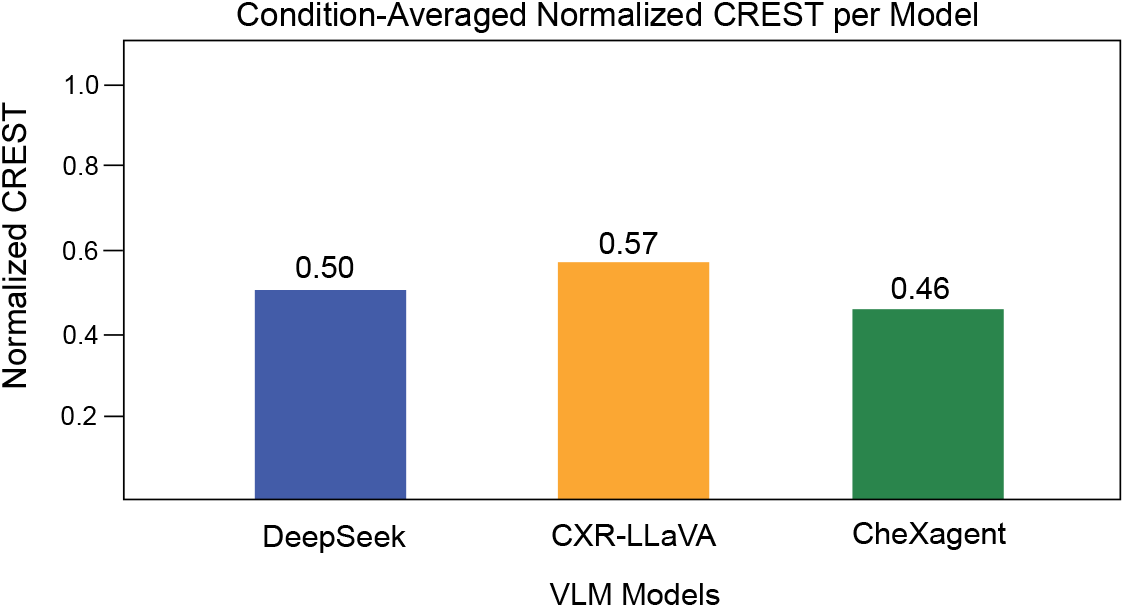
Condition-Averaged Clinical Risk-weighted Error Score for Text-generation (CREST) of VLMs.

Figure 6 presents the normalized CREST per condition across the 13 clinical categories. CheXagent achieves lower scores in high-risk conditions such as Pneumothorax, Pneumonia, and Atelectasis, suggesting it tends to avoid high-severity mistakes in these areas. Interestingly, DeepSeek outperforms CXR-LLaVA in several conditions, including some high-risk ones (e.g. Pneumonia). This suggests that its general-purpose training may help mitigate certain types of clinically impactful mistakes, despite the lack of radiology-specific fine-tuning.

**Figure 6:**
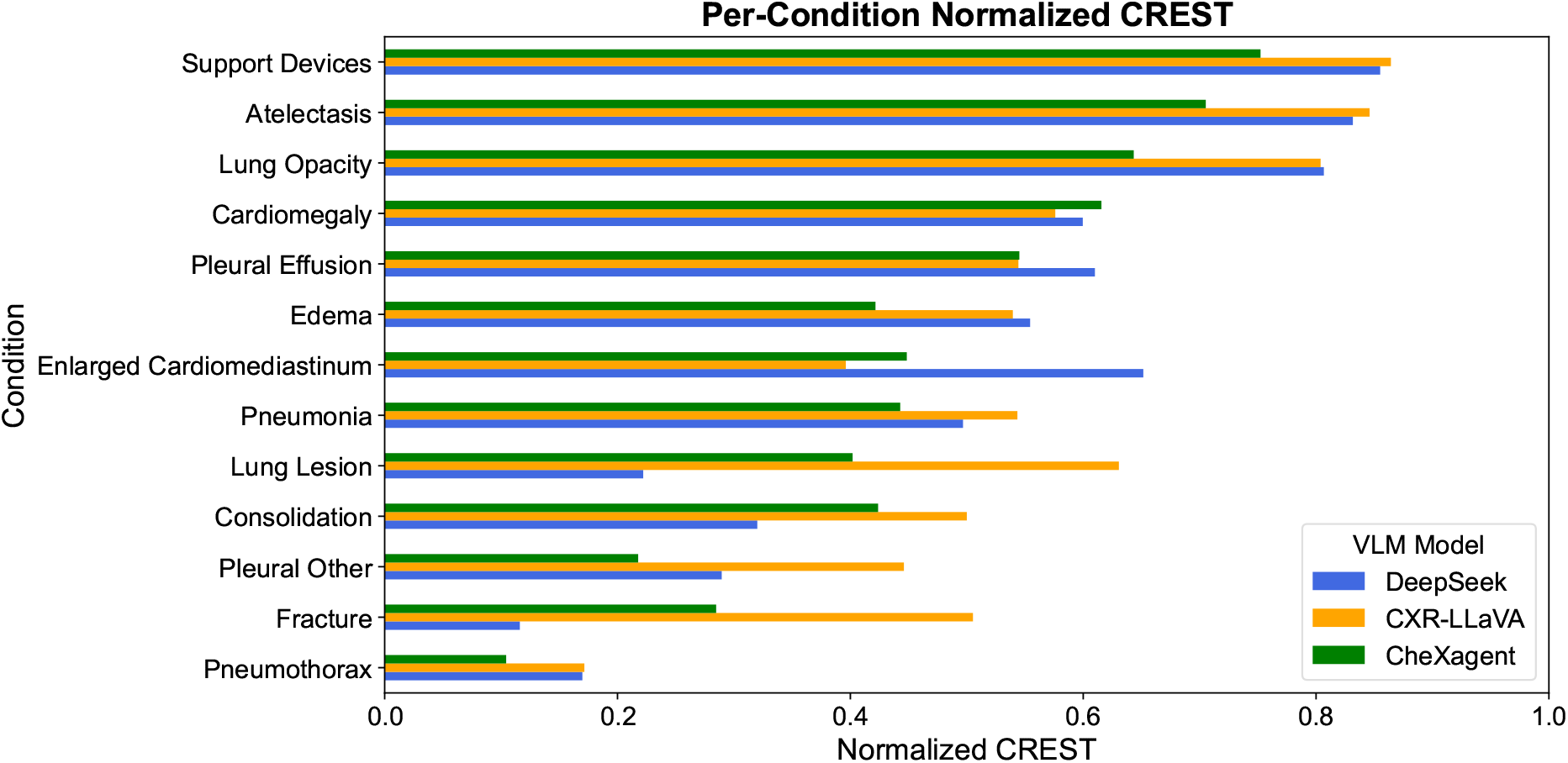
Conditional-Level Clinical Risk-weighted Error Score for Text-generation (CREST) of VLMs. of AI-generated content.

The conditon-averaged CREST provides a useful risk summary, while the condition-level scores offer more detailed guidance for model evaluation and refinement. Together, these perspectives demonstrate CREST’s advantage over traditional metrics by aligning model evaluation with clinical relevance.

## Discussion and Conclusions

While our study compares three representative VLMs, the primary contribution (focus) is the development and demonstration of CREST, a novel evaluation framework (including a taxonomy of errors that captures the range and nature of clinically relevant mistakes) designed to quantify the clinical severity of errors made by VLMs in radiology report generation. The comparison of VLMs serves to illustrate how CREST can uncover clinically meaningful differences in model behavior beyond traditional metrics. Our results reveal that models with similar overall error counts can differ significantly in clinical risk, highlighting the importance of evaluating not just the quantity, but also the impact

Unlike traditional evaluation metrics that focus on semantic similarity or binary correctness, CREST accounts for the clinical significance of each error, offering a more meaningful and task-relevant assessment. This framework is highly generalizable: while demonstrated in radiology report generation, it can be extended to other medical text-generation tasks such as discharge summaries or clinical decision support outputs where the consequences of model outputs carry varying levels of risk. Importantly, CREST can be automatically computed, avoiding the need for extensive human expert annotation that is often required for clinical validation.

Looking ahead, risk-aware metrics like CREST open new avenues for detecting and analyzing hallucinations, omissions, and biases in generative models. By quantifying the potential harm of individual errors, these metrics can help prioritize failure cases, guide fine-tuning, and inform post-processing or retrieval-augmented generation strategies aimed at minimizing clinically significant mistakes.

The current version of CREST has several limitations. 1) It uses fixed weights for clinical risk levels (i.e., Low = 1, Medium = 2, High = 3), which may lack flexibility across diverse clinical settings. Future work may adopt a decision-theoretic utility function that reflects real-world clinical costs, varying by setting, country, task, or patient history. 2) The impact of prompt variation on model performance is not explored and will be addressed in future work. 3) CREST aggregates risk-weighted errors independently across conditions and does not account for comorbidities. Future extensions may incorporate comorbidity modeling. 4) The number of evaluated cases is relatively small, and annotations were performed by a single expert, which may introduce bias. Future work will expand the dataset and include multiple annotators. We acknowledge that this study represents an initial implementation of the CREST metric. While grounded in clinical expert consensus, formal validation of its reliability and utility remains an important direction for future research.

## Data Availability

All data analyzed in the present study are publicly available from PhysioNet at https://physionet.org upon completion of credentialing and data use agreement.

## Acknowledgments

This research was funded by NIH/NLM 1R01LM014239.

## Notes

### Competing Interest Statement

The authors have declared no competing interest.

### Author Declarations

The Institutional Review Board of the Massachusetts Institute of Technology waived ethical approval for this work.

## References

1. Zhang J, Huang J, Jin S, Lu S. Vision-language models for vision tasks: A survey. IEEE Transactions on Pattern Analysis and Machine Intelligence. 2024.

2. Hartsock I, Rasool G. Vision-language models for medical report generation and visual question answering: A review. Frontiers in Artificial Intelligence. 2024;7:1–26.

3. Yi Z, Xiao T, Albert MV. A Survey on Multimodal Large Language Models in Radiology for Report Generation and Visual Question Answering. Information. 2025;16(2):1–27.

4. Reale-Nosei G, Amador-Domínguez E, Serrano E. From vision to text: A comprehensive review of natural image captioning in medical diagnosis and radiology report generation. Medical Image Analysis. 2024:1–20.

5. Tanno R, Barrett DG, Sellergren A, Ghaisas S, Dathathri S, See A, et al. Collaboration between clinicians and vision–language models in radiology report generation. Nature Medicine. 2025;31(2):599–608.

6. Monshi MMA, Poon J, Chung V. Deep learning in generating radiology reports: A survey. Artificial Intelligence in Medicine. 2020;106:1–12.

7. Kaur N, Mittal A, Singh G. Methods for automatic generation of radiological reports of chest radiographs: a comprehensive survey. Multimedia Tools and Applications. 2022;81(10):13409–39.

8. Johnson AE, Pollard TJ, Berkowitz SJ, Greenbaum NR, Lungren MP, Deng Cy, et al. MIMIC-CXR, a de-identified publicly available database of chest radiographs with free-text reports. Scientific Data. 2019;6(1):1–8.

9. Johnson AE, Pollard TJ, Greenbaum NR, Lungren MP, Deng Cy, Peng Y, et al. MIMIC-CXR-JPG, a large publicly available database of labeled chest radiographs. arXiv preprint arXiv:190107042. 2019.

10. Wu Z, Chen X, Pan Z, Liu X, Liu W, Dai D, et al. Deepseek-vl2: Mixture-of-experts vision-language models for advanced multimodal understanding. arXiv preprint arXiv:241210302. 2024.

11. Lu H, Liu W, Zhang B, Wang B, Dong K, Liu B, et al. Deepseek-vl: towards real-world vision-language understanding. arXiv preprint arXiv:240305525. 2024.

12. Lee S, Youn J, Kim H, Kim M, Yoon SH. CXR-LLAVA: a multimodal large language model for interpreting chest X-ray images. European Radiology. 2025:1–13.

13. Liu H, Li C, Wu Q, Lee YJ. Visual instruction tuning. Advances in Neural Information Processing Systems. 2023;36:34892–916.

14. Chen Z, Varma M, Delbrouck JB, Paschali M, Blankemeier L, Van Veen D, et al. CheXagent: Towards a Foundation Model for Chest X-Ray Interpretation. In: AAAI 2024 Spring Symposium on Clinical Foundation Models; 2024. p. 1–4.

15. Smit A, Jain S, Rajpurkar P, Pareek A, Ng AY, Lungren M. Combining Automatic Labelers and Expert Annotations for Accurate Radiology Report Labeling Using BERT. In: Proceedings of the 2020 Conference on Empirical Methods in Natural Language Processing (EMNLP); 2020. p. 1500–19.

16. Bluethgen C, Chambon P, Delbrouck JB, van der Sluijs R, Połacin M, Zambrano Chaves JM, et al. A vision– language foundation model for the generation of realistic chest x-ray images. Nature Biomedical Engineering. 2024:1–13.

17. Ji J, Hou Y, Chen X, Pan Y, Xiang Y. Vision-language model for generating textual descriptions from clinical images: Model development and validation study. JMIR Formative Research. 2024;8:1–13.

18. Endo M, Krishnan R, Krishna V, Ng AY, Rajpurkar P. Retrieval-based chest x-ray report generation using a pre-trained contrastive language-image model. In: Machine Learning for Health. PMLR; 2021. p. 209–19.

19. Srivastav S, Ranjit M, Pérez-García F, Bouzid K, Bannur S, Castro DC, et al. MAIRA at RRG24: A specialised large multimodal model for radiology report generation. In: Proceedings of the 23rd Workshop on Biomedical Natural Language Processing; 2024. p. 597–602.

